# Distribution and Patterns of Device-Measured Movement Behaviours in middle-aged to older Australian adults: the ABC Accelerometer Sub-Study

**DOI:** 10.64898/2026.07.26.26358971

**Authors:** Jay Keatley, Nga Nguyen, Paddy C. Dempsey, Simone J.J.M. Verswijveren, Julie K. Bassett, Roger L. Milne, Brigid M. Lynch

## Abstract

**Objectives:** To describe device-measured movement behaviours in a large sample of middle-aged and older Australian adults using complementary posture- and intensity-based accelerometers, examine variation across demographic groups, and identify behavioural phenotypes using clustering approaches

**Design:** Cross-sectional analysis of the Australian Breakthrough Cancer (ABC) Study Accelerometer Sub-study (ACM).

**Methods:** Participants from the ABC cohort completed seven days of simultaneous monitoring using a thigh-mounted activPAL and waist-mounted ActiGraph GT3X+. activPAL characterised posture-based behaviours (sitting, lying, standing, stepping, postural transitions, and sedentary accumulation), while ActiGraph characterised intensity-based activity (sedentary, light, and moderate-to-vigorous physical activity [MVPA]). Movement behaviours were summarised overall and by gender, age group, body mass index (BMI), and education. Behavioural phenotypes were identified using k-means clustering.

**Results:** Among 4,238 ACM participants, 3,426 met inclusion criteria with valid data from both devices (1,711 females; 1,715 males). Participants accumulated substantially more time in sedentary and low-intensity behaviours than in MVPA. activPAL estimates indicated mean daily time of 379.3 min sitting, 282.2 min lying, 160.8 min standing, and 64.5 min stepping, with a mean of 5,185 steps/day. ActiGraph estimates indicated 584.0 min/day sedentary time, 290.9 min/day light-intensity activity, and 33.2 min/day MVPA. Considerable heterogeneity in movement behaviours was observed between individuals, whereas demographic differences were comparatively modest. Three behavioural phenotypes were identified: active/fragmented (24%), low activity (41%), and prolonged sedentary (35%). Notably, the low-activity and prolonged sedentary phenotypes were distinct, indicating that low overall movement and prolonged uninterrupted sitting represented different behavioural patterns. The prolonged sedentary phenotype was characterised by greater uninterrupted sitting time, lower stepping time, fewer steps, lower MVPA, and fewer sit-to-stand transitions.

**Conclusions:** Movement behaviours in middle-aged to older Australian adults (40-74 yrs) were characterised by high sedentary time, low accumulation of MVPA, and substantial between-person heterogeneity. Distinct behavioural phenotypes highlighted differences in both movement volume and sedentary accumulation patterns, suggesting that movement behaviour is multidimensional and not adequately described by single summary measures alone. These findings may help inform our understanding of population movement patterns relevant to cancer and cardiometabolic disease prevention.

## Introduction

Movement behaviours (lying, sitting, standing, and physical activity) are increasingly recognised as important determinants of chronic disease risk and healthy ageing. Higher levels of physical activity are associated with lower risks of several cancers, cardiovascular disease, type 2 diabetes, and premature mortality, whereas prolonged sedentary behaviour has been linked with adverse health outcomes independent of physical activity levels (Ekelund et al., 2019; Matthews et al., 2020; Moore et al., 2016). Contemporary public health guidelines therefore emphasise not only increasing physical activity but also reducing and interrupting sedentary time. Understanding how movement behaviours are distributed and accumulated within populations is important for identifying behavioural patterns relevant to disease prevention, informing public health recommendations, and establishing baseline population profiles against which future health outcomes can be studied.

Large, cohort representative descriptive data can characterise how movement behaviours are distributed within communities and identify behavioural patterns relevant to health outcomes. Historically, cohort studies assessing movement behaviours have relied on self-reported questionnaires or interviews. Although these tools provide important contextual information, they are prone to recall bias, social desirability, and misclassification (Prince et al., 2010; Helmerhorst et al., 2012), particularly for low-intensity and incidental activities that dominate daily behaviour. Self-reported measures are known to underestimate sedentary behaviour and misclassify activity intensity compared with accelerometry (Prince et al., 2010; Steene-Johannessen et al., 2016).

Accelerometry enables objective measurement of posture and physical activity intensity. Accelerometers can quantify sitting, standing, stepping, sedentary accumulation, postural transitions, cadence, and activity intensity, providing a more detailed characterisation of movement behaviours than self-report measures (Helbostad et al., 2016; Prince et al., 2010). Few studies have described device-measured movement behaviours in large samples, particularly within an Australian context. In addition, limited evidence exists regarding heterogeneity in the way individuals accumulate movement behaviours. Beyond traditional descriptive summaries, data-driven approaches such as clustering algorithms can identify distinct behavioural phenotypes, providing insight into variation in movement behaviour patterns within populations.

Most large cohort studies have relied on a single accelerometer to characterise movement behaviours. However, posture-based and intensity-based accelerometers capture different dimensions of behaviour and are not directly interchangeable. The activPAL provides posture-based measures of sitting/lying, standing, stepping, and sedentary accumulation, whereas the ActiGraph provides intensity-based measures of sedentary, light-, moderate-, and vigorous-intensity physical activity. Concurrent use of both devices enables a more comprehensive assessment of movement behaviours by combining information on both postural allocation and activity intensity.

The Australian Breakthrough Cancer (ABC) Study Accelerometer Sub-study (ACM) provides a unique opportunity to characterise device-measured movement behaviours in a large sample of middle-aged and older Australian adults using complementary posture- and intensity-based accelerometers. The aims of this study were to: (1) describe the distribution of activPAL- and ActiGraph-derived movement behaviours; (2) examine variation in movement behaviours according to gender, age, body mass index, and educational attainment; and (3) identify behavioural phenotypes using clustering approaches to characterise heterogeneity in movement behaviours.

## Methods

The Australian Breakthrough Cancer Study is a large cohort of Australian adults recruited between 2014 and 2018, with 56,282 participants completing the baseline questionnaires. The ABC Study was established to investigate the role that genetics, lifestyle and environment play in the development of cancer and other diseases.

The ABC Accelerometer Sub-study was conducted between 2016 and 2020 as a nested pilot designed to evaluate the feasibility and acceptability of assessing movement behaviours by accelerometry. Participants were eligible if they had completed the ABC Study baseline questionnaires and had been invited to provide a blood sample. Participants provided written informed consent and were mailed accelerometers to be used for seven consecutive days of monitoring. The ABC Study has been approved by the Human Research Ethics Committee of Cancer Council Victoria. This study has been carried out according to the National Statement on Ethical Conduct in Human Research (2007).

### Accelerometer assessment

Participants wore two triaxial accelerometers simultaneously: a thigh-mounted activPAL (PAL Technologies, Glasgow, UK) and a waist-mounted ActiGraph GT3X+ (ActiGraph LLC, Pensacola, FL, USA). The activPAL was attached to the anterior midline of the thigh using adhesive dressing and worn continuously for 24 hours per day, including during sleep. The ActiGraph was worn on an elastic belt at the mid-axillary line during waking hours and removed during sleep and water-based activities. Participants completed wear diaries documenting sleep and device removal periods.

activPAL valid days were defined as 24-hour recordings with <4 hours of non-wear. activPAL data were processed using proprietary software that classifies behaviour based on thigh inclination and acceleration. Derived variables included sitting time, standing time, stepping time, lying time, upright time, and recumbent posture metrics. Static upright periods were classified as standing, whereas dynamic upright periods were classified as stepping. Non-upright static periods were classified as sitting/lying. Lying time was derived from sustained non-upright posture which may include sleep as well as waking rest and should therefore be interpreted as time spent in a recumbent posture rather than sleep per se. Additional activPAL-derived measures included daily step counts, sit-to-stand transitions, prolonged sitting bouts (≥30 and ≥60 min), time accumulated in prolonged sitting bouts (≥30 and ≥60 min), and cadence-based stepping measures (≥75 and ≥100 steps/min). Daily summaries were generated for all valid wear periods and averaged across valid monitoring days for each participant.

ActiGraph valid days were defined as ≥8 hours of wear time. Non-wear periods were identified during data processing and days with wear time <480 minutes were excluded. ActiGraph-derived outcomes included sedentary behaviour, light-intensity physical activity, moderate-intensity physical activity, vigorous-intensity physical activity, and moderate-to-vigorous physical activity (MVPA). Activity intensities were classified using the Freedson adult cut-points, with sedentary behaviour defined as <100 counts/min, light activity as 100–1951 counts/min, moderate activity as 1952–5724 counts/min, and vigorous activity as ≥5725 counts/min (Freedson et al., 1998). MVPA was calculated as the sum of moderate- and vigorous-intensity activity. Daily summaries were generated for all valid wear periods and averaged across valid monitoring days for each participant. Processed outputs from both devices were merged using participant identifiers and calendar date. All data management and analyses were conducted in RStudio version 2024.12.0 (Posit Software, PBC 2024).

### Valid wear criteria and data cleaning

Device-specific quality-control procedures were applied independently to activPAL and ActiGraph data prior to analysis. Daily records with insufficient wear time, implausible behaviour totals, excessive vector magnitude values, or device error flags were excluded. Participant-level summaries were derived using valid days only. A minimum threshold of ≥1 valid wear day for both devices was used for inclusion to maximise retention and support descriptive characterisation of movement behaviours across the cohort. The final analytic sample included participants with valid data from both devices after quality control. Sensitivity analyses examined alternative wear-time thresholds and exclusion of extreme values. Detailed quality-control procedures and thresholds are provided in the Supplementary Methods.

### Statistical analysis

Movement behaviours were summarised using means, standard deviations, medians, and interquartile ranges. Behavioural variables were expressed as minutes per day, except step counts and sit-to-stand transitions, which were expressed as counts per day. Estimates were presented overall and stratified by gender, age group, BMI category, and education. Gender was derived from self-reported questionnaire data. Participants identifying as transgender were classified according to their reported gender identity (transgender women as female and transgender men as male). No separate gender-diverse category was available for analysis due to small numbers. As the ABC Study collected self-reported gender identity rather than biological sex assigned at birth, gender was used throughout the present analyses and manuscript. We also used violin plots to demonstrate the distribution of movement behaviours. Behavioural phenotypes were identified using k-means clustering, an unsupervised partitioning method that groups individuals with similar multivariable movement behaviour profiles into mutually exclusive clusters while minimising within-cluster variation and maximising between-cluster differences. Input variables comprised standardised measures of overall movement volume and behavioural accumulation patterns, including lying time, sedentary time, standing time, stepping time, daily steps, sit-to-stand transitions, and prolonged sedentary bout metrics. All variables were converted to z-scores prior to analysis to ensure comparability across different measurement scales and to prevent variables with larger numeric ranges disproportionately influencing cluster allocation. Candidate cluster solutions (k = 2 to 6) were examined and compared according to cluster size, profile separation, interpretability, and epidemiological relevance. A three-cluster solution was selected as the most parsimonious and meaningful representation of the data, balancing distinct behavioural patterns with adequate cluster sizes for interpretation. Cluster solutions were evaluated based on interpretability and size and visualised using heatmaps and radar plots. Sensitivity analyses examined alternative wear-time thresholds and exclusion of extreme values.

## Results

From the 4,238 participants in ACM 3,426 met the primary inclusion criterion of at least one valid wear day for both devices. Participants with insufficient valid wear time, incomplete device returns, or records failing quality-control procedures were excluded. The analytic sample comprised 1,711 females and 1,715 males. Device wear-time compliance was high. For activPAL, all included participants contributed at least one valid day for both devices, and 99.7% of monitored days achieved complete 24-hour recording. For ActiGraph, the median daily wear time was 894 minutes (14.9 h/day) (IQR 840–949).

Females were slightly younger than males (mean age 60.3 vs 61.2 years), although the age distribution was broadly similar between genders, with most participants aged 50–69 years. Mean BMI was comparable between females and males, although overweight was more common among males, while females were more frequently in the healthy weight BMI range. Obesity prevalence was similar between genders. Approximately half of participants reported a university degree or higher, with broadly similar educational attainment across gender (Table 1).

**Table 1.** Participant demographic characteristics (N = 3,425)

| Characteristic | Female (N = 1,710) | Male (N = 1,715) |
| --- | --- | --- |
| **Age, years** | 60.3 ± 8.2 | 61.2 ± 8.3 |
| **Age group, years** |  |  |
| 40–49 | 206 (12%) | 192 (11%) |
| 50–59 | 584 (34%) | 520 (30%) |
| 60–69 | 704 (41%) | 731 (43%) |
| 70–80 | 213 (12%) | 271 (16%) |
| **Body mass index, kg/m<sup>2</sup>** | 27.0 ± 6.0 | 27.5 ± 4.9 |
| **Body mass index group** |  |  |
| Normal (18.5–24.9) | 671 (39%) | 490 (29%) |
| Overweight (25–29.9) | 564 (33%) | 784 (46%) |
| Obese (≥30) | 417 (24%) | 392 (23%) |
| Underweight (<18.5) | 21 (1%) | 13 (1%) |
| Missing | 37 (2%) | 36 (2%) |
| **Education** |  |  |
| Certificate/diploma | 447 (26%) | 529 (31%) |
| Junior secondary | 258 (15%) | 152 (9%) |
| Primary education or less | 9 (1%) | 11 (1%) |
| Senior secondary education | 185 (11%) | 161 (9%) |
| University degree or higher | 804 (47%) | 849 (50%) |
| Missing | 7 (0%) | 13 (1%) |

*Values are presented as mean ± standard deviation (SD) for continuous variables and n (%) for categorical variables.*

### activPAL results

activPAL data indicated that participants accumulated substantially more time lying or sitting than standing or stepping (Table 2). Mean daily time was 379.3 min sitting, 282.2 min lying, 160.8 min standing, and 64.5 min stepping, indicating that sitting and lying together accounted for approximately 11 hours/day of recorded posture-based behaviour. Mean daily step count was 5,185 steps/day (SD 2,359).

**Table 2.**
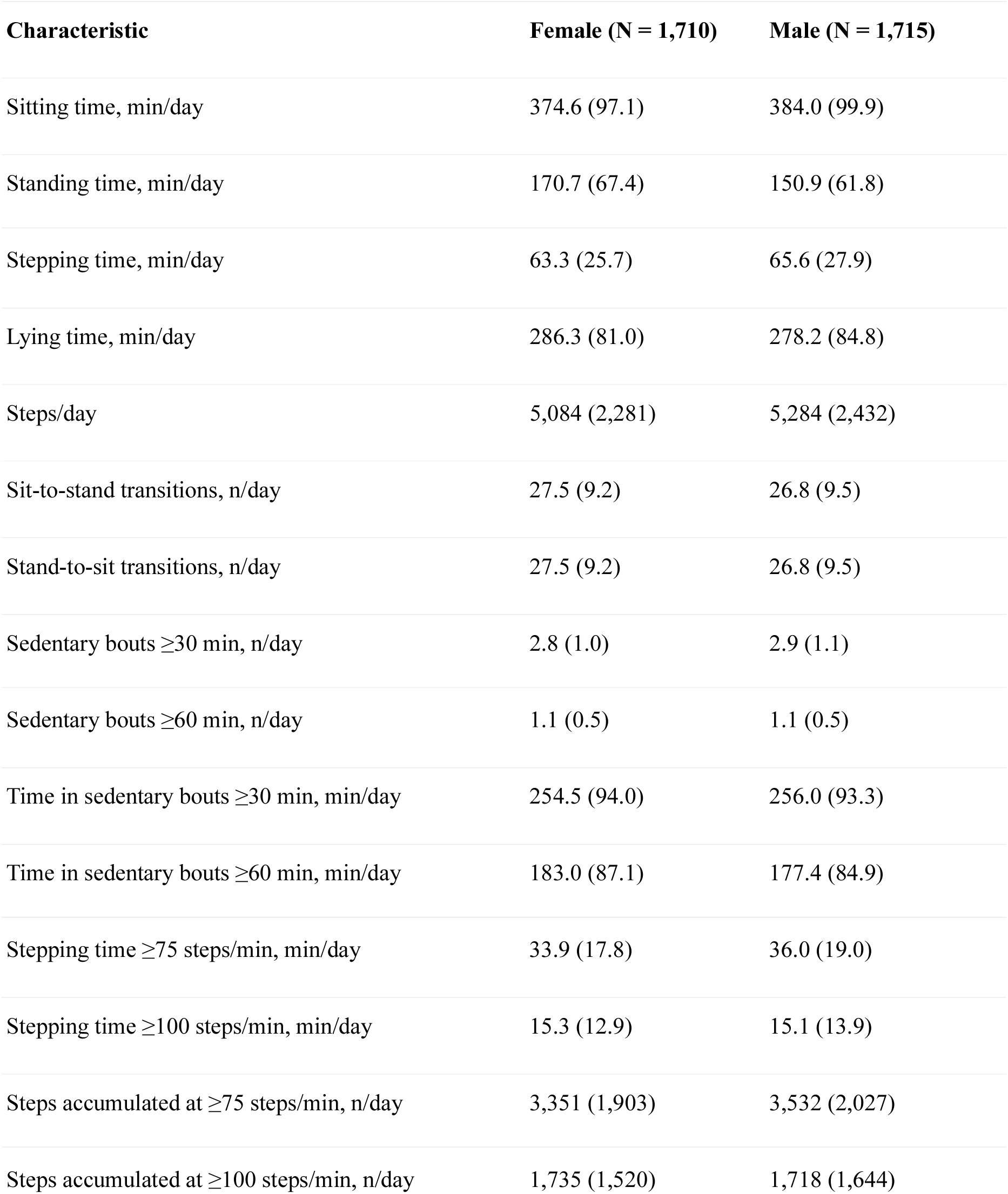
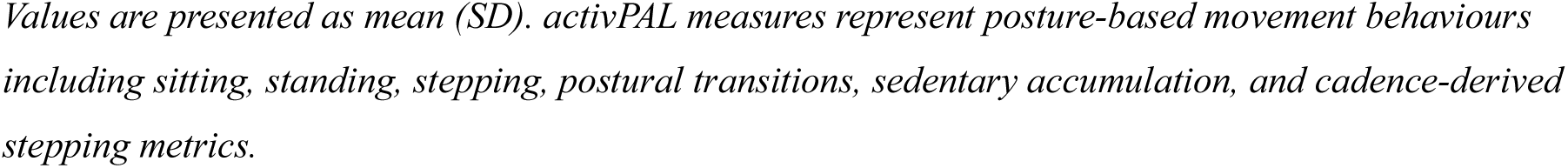
Posture-based daily movement behaviours measured by activPAL, by gender.

Gender differences were modest. Males accumulated slightly more sitting time (384.0 vs 374.6 min/day) and stepping time (65.6 vs 63.3 min/day), whereas females accumulated more standing time (170.7 vs 150.9 min/day) and lying time (286.3 vs 278.2 min/day) (Table 2).

Daily step counts were modest and right-skewed, with most participants accumulating approximately 4,000–7,000 steps/day and a smaller subgroup achieving substantially higher values. Cadence was relatively consistent (∼70–85 steps/min), indicating most stepping occurred at light-to-moderate intensity (Figure 2a). Across gender, age group, BMI category, and education groups, cadence was broadly similar, whereas step counts were generally lower among older adults, participants with obesity, and those with lower educational attainment (Figure 2b; Supplementary Figure 2a–c).

**Figure 1.**
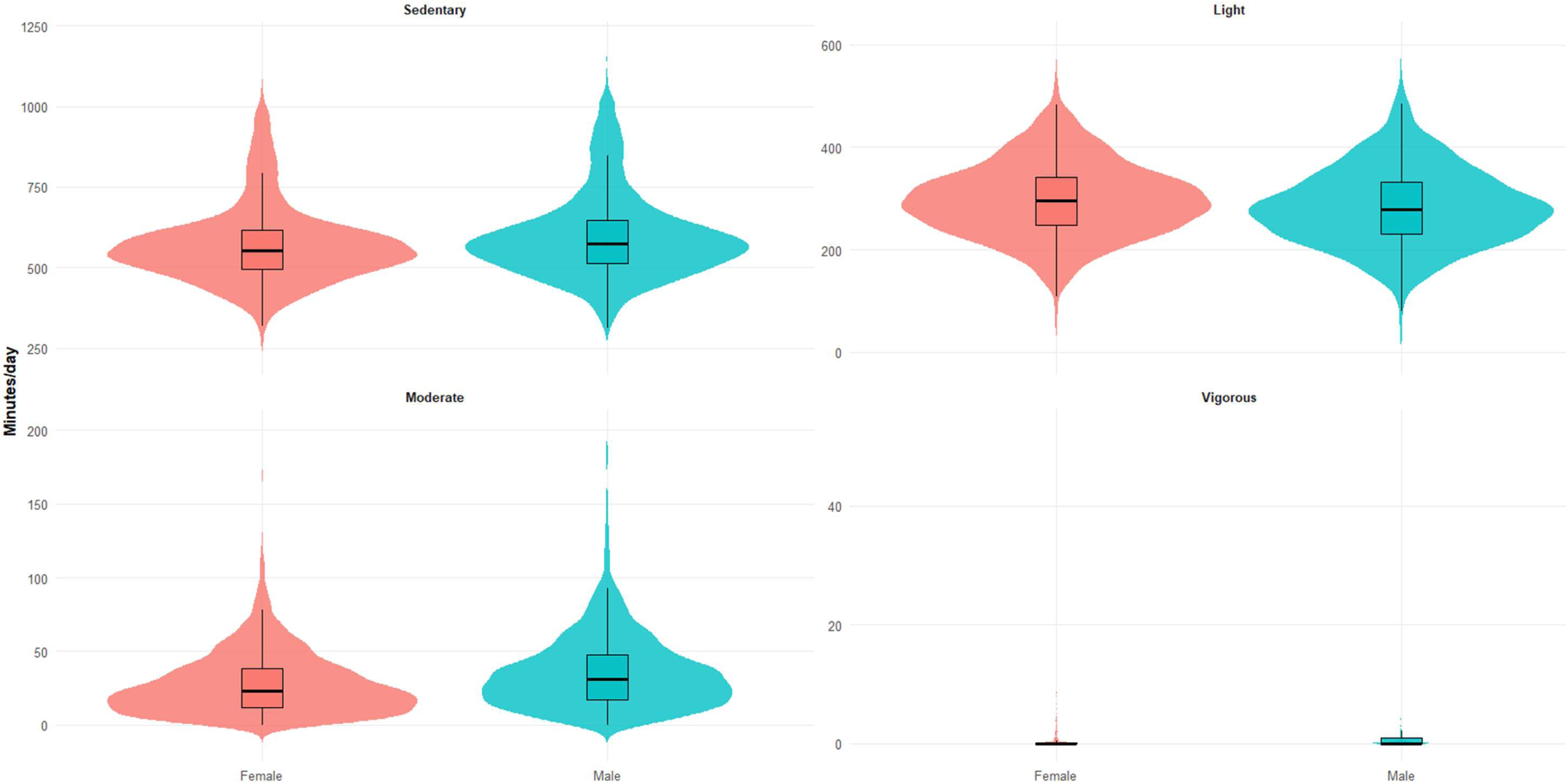
Distribution of Actigraph-derived activity intensities, by gender.

**Figure 2a.**
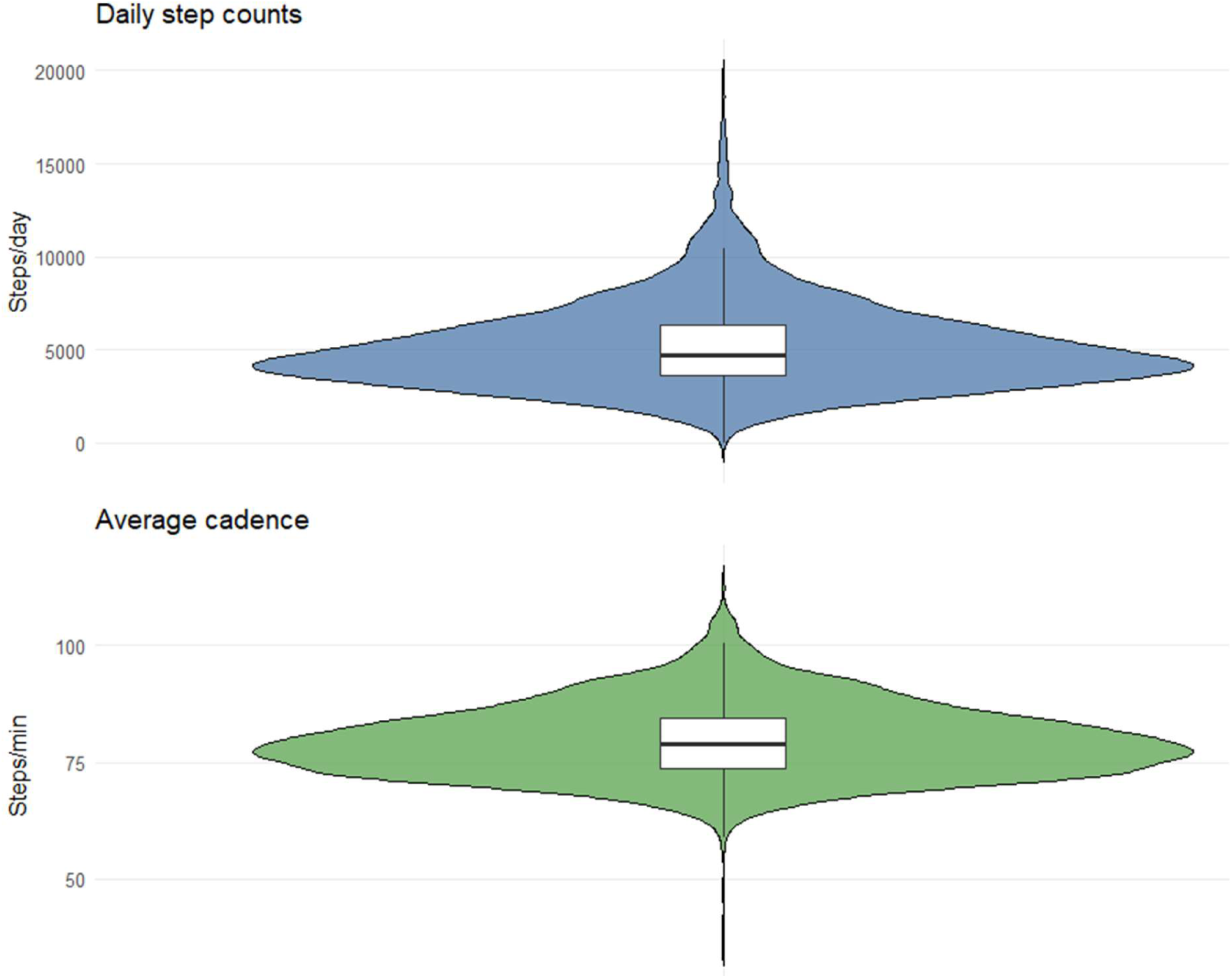
Distribution of step counts and cadence, overall

**Figure 2b.**
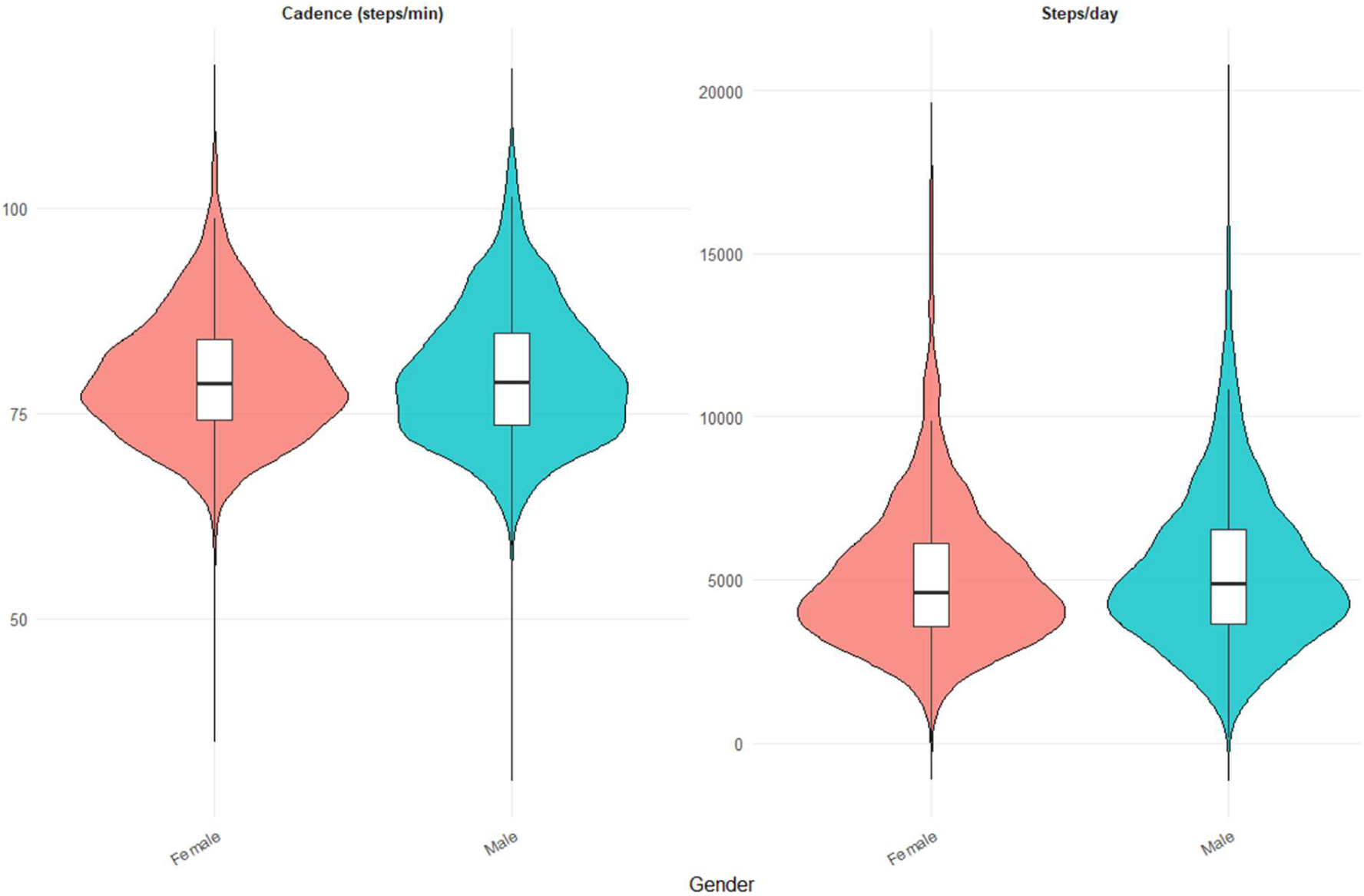
Distribution of step counts and cadence, by gender

Consistent difference across BMI and age groups were observed. Participants with obesity accumulated lower stepping time (55.4 vs 70.5 min/day) and fewer daily steps (4,346 vs 5,787 steps/day) than those in the healthy weight BMI range. Older adults, particularly those aged 70–80 years, also accumulated fewer daily steps and slightly higher sitting time than younger participants (Supplementary Table 1a–b).

Participants recorded approximately 27 sit-to-stand transitions per day, indicating moderate fragmentation of sedentary behaviour (Table 2). On average, they accumulated 2.9 sedentary bouts/day of ≥30 minutes and 1.1 bouts/day of ≥60 minutes. A large proportion of sitting time was accumulated in prolonged bouts, indicating that sedentary behaviour was not only high in volume but also frequently accumulated in extended, uninterrupted periods. This included 255.2 min/day in bouts ≥30 minutes and 180.2 min/day in bouts ≥60 minutes

Patterns were broadly similar between females and males, although females recorded slightly more daily postural transitions and lower total sitting time (Table 2). Older adults and participants with obesity accumulated fewer sit-to-stand transitions, more sedentary bouts, and greater time in prolonged sitting bouts than younger or healthy weight participants. Differences by education were small (Supplementary Table 2a–c).

### ActiGraph results

Consistent with activPAL findings, ActiGraph estimates indicated that participants spent the majority of the day sedentary or in light-intensity activity, with only a small proportion of time in MVPA (Table 3). Mean daily time was 584.0 min sedentary, 290.9 min light-intensity activity, and 33.2 min MVPA, with minimal vigorous activity overall. Sedentary and light-intensity activity together accounted for approximately 14.5 hours/day.

**Table 3.** Intensity-based daily movement behaviours measured by ActiGraph, by gender.

| Characteristic | Female (N = 1,710) | Male (N = 1,715) |
| --- | --- | --- |
| Sedentary time, min/day | 570.8 (121.2) | 597.1 (132.9) |
| Light-intensity activity, min/day | 297.5 (72.4) | 283.4 (75.5) |
| Moderate-intensity activity, min/day | 27.6 (20.7) | 35.6 (25.2) |
| Vigorous-intensity activity, min/day | 1.0 (3.1) | 2.0 (5.5) |
| MVPA, min/day | 28.6 (21.8) | 37.6 (27.0) |
| Steps/day | 6,842 (5,173–9,173) | 7,562 (5,474–9,801) |

*Values are presented as mean (SD), except steps/day which are presented as median (IQR). MVPA = moderate-to-vigorous physical activity. ActiGraph measures represent intensity-based movement behaviours derived from accelerometry.*

Gender differences were modest but consistent. Males accumulated more MVPA (37.6 vs 28.6 min/day), whereas females accumulated more light-intensity activity (297.5 vs 283.4 min/day) (Figure 1).

Although some demographic gradients were evident, particularly for BMI and age, substantial overlap between groups was observed across most movement behaviours. Participants with obesity accumulated less MVPA (24.0 vs 39.3 min/day) and fewer daily steps (5,833 vs 8,202 steps/day) than those in the normal BMI range. Older adults, particularly those aged 70–80 years, also tended to accumulate fewer daily steps, less moderate activity, and lower MVPA than younger participants. Participants with lower educational attainment similarly recorded fewer steps and lower moderate-intensity activity than those with higher education levels (Supplementary Table 1a–c).

### Behavioural phenotypes

K-means clustering identified three behavioural phenotypes based on activity volume, intensity, and sedentary accumulation (Figure 3; Supplementary Table 4). The three-cluster solution was selected based on interpretability and balanced cluster sizes, as solutions with more clusters resulted in small groups that were difficult to interpret.

**Figure 3.**
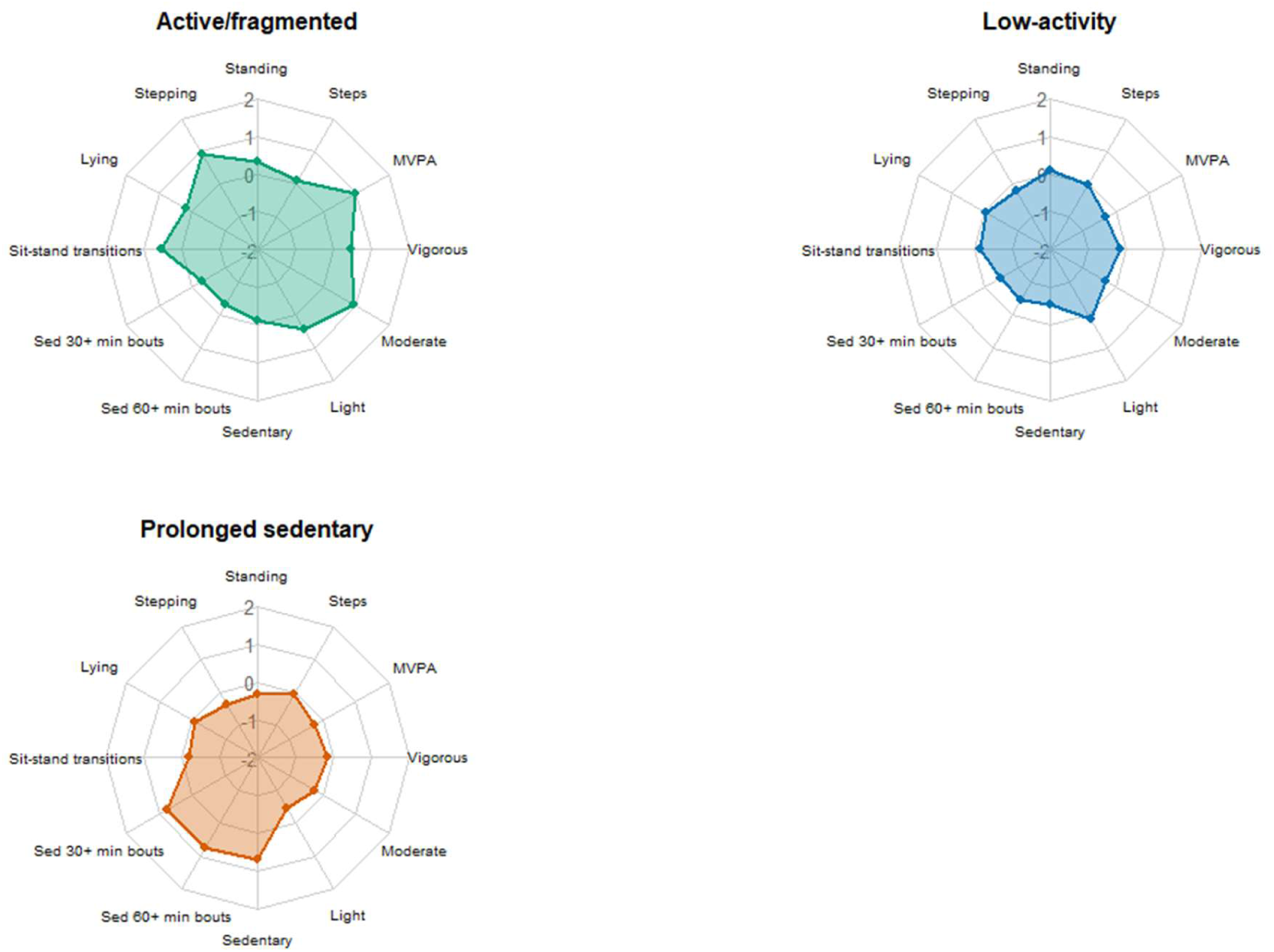
Radar plots of behavioural phenotypes Values are z-scores relative to the cohort mean, where 0 represents the sample average. Positive values indicate higher-than-average values and negative values indicate lower-than-average values for each movement characteristic. Points further from the centre indicate relatively higher values. Active/Fragmented phenotype (Cluster 1) shows higher steps/day, MVPA, stepping, and sit-to-stand transitions. Prolonged Sedentary phenotype (Cluster 3) shows higher sedentary time and prolonged sedentary bout accumulation, consistent with a prolonged sedentary profile. Absolute values are presented in Supplementary Table 4.

The low-activity (Cluster 2) and prolonged sedentary (Cluster 3) phenotypes were distinct, indicating that low overall movement and prolonged uninterrupted sitting represented different behavioural patterns. The active/fragmented phenotype (Cluster 1) (n = 804; 24%) had the highest levels of steps, MVPA, and stepping time (8,020.3 steps/day, 63.3 min/day MVPA, and 95.7 min/day stepping), along with more frequent sit-to-stand transitions (33.4/day) and less prolonged sedentary time (147.4 min/day in bouts ≥60 minutes). The low-activity phenotype (n = 1,409; 41%) was characterised by consistently low activity levels (4,505.8 steps/day, 23.4 min/day MVPA) and fewer transitions (25.6/day), without clearly elevated prolonged sedentary time (129.5 min/day in bouts ≥60 minutes). The prolonged sedentary phenotype (n = 1,215; 35%) had the highest accumulation of sedentary time (260.5 min/day in bouts ≥60 minutes), alongside low stepping time (51.3 min/day), lower steps (4,096.2 steps/day), low MVPA (24.4 min/day), and fewer transitions (24.7/day).

A heatmap further illustrates these differences, with the active phenotype showing higher activity across all intensity measures, while the prolonged sedentary phenotype was characterised by higher sedentary accumulation and lower movement (Supplementary Figure 3). These results indicate clear differences in how movement behaviours were accumulated across individuals.

## Discussion

In this large cross-sectional study of middle-aged and older Australian adults, device-measured movement behaviours were characterised by a predominance of sedentary behaviour and light-intensity activity, with only a small proportion of time spent in MVPA. This pattern is consistent with findings from other large accelerometer-based cohort studies. For example, the UK Biobank and China Kadoorie Biobank similarly demonstrate that free-living activity is dominated by lower-intensity movement, with relatively little time accumulated at higher intensities (Doherty et al., 2017; Chen et al., 2023). Together, these findings suggest that the predominance of sedentary and light-intensity behaviours represents a consistent pattern. The present study extends this literature by providing one of the largest descriptions of device-measured movement behaviours in an Australian cohort using complementary posture- and intensity-based accelerometers.

A key finding from our study was the substantial heterogeneity in movement behaviours between individuals. Differences in sedentary time, step counts, and MVPA were substantially larger between participants than between demographic groups defined by gender, age, BMI, or educational attainment. Although older adults and individuals with higher BMI tended to accumulate more sedentary time and had lower activity levels than others, these differences were small relative to the wide variability observed within the whole cohort. Similar heterogeneity in objectively measured physical activity has been reported in other accelerometer studies, where substantial overlap exists between demographic groups despite differences in average activity levels (Golubic et al., 2014; Doherty et al., 2017).

Sedentary behaviour was characterised by prolonged accumulation, with a substantial proportion of sitting time occurring in uninterrupted bouts. Similar patterns of prolonged sedentary accumulation have been observed in previous activPAL-based studies of middle-aged and older adults (van der Berg et al., 2016). Consistent with these observations, a large proportion of sitting time in the present cohort was accumulated in bouts lasting ≥30 and ≥60 minutes. Overall accumulation of MVPA was low, with most participants recording relatively limited higher-intensity activity. The use of complementary device-based measures provides additional insight into movement behaviours by capturing both posture (activPAL) and activity intensity (ActiGraph). This distinction is increasingly recognised as important, as sedentary behaviour and physical inactivity represent related but distinct constructs, and posture-based and intensity-based measures capture complementary dimensions of movement behaviour (Dempsey et al., 2020).

Differences between activPAL and ActiGraph estimates were expected and reflect the distinct behavioural constructs measured by each device. activPAL primarily characterises posture-based behaviours using thigh inclination, allowing differentiation of sitting/lying, standing, and stepping, whereas ActiGraph characterises movement intensity based on acceleration-derived activity counts. Consequently, ActiGraph estimated higher sedentary time than activPAL, likely reflecting the classification of low-movement standing behaviours as sedentary under intensity-based thresholds.

Finally, clustering analyses identified three distinct behavioural phenotypes: active/fragmented, low-activity, and prolonged sedentary, highlighting that movement behaviour is multidimensional and not adequately described by summary metrics alone. Notably, the low-activity and prolonged sedentary phenotypes emerged as separate profiles, indicating that low overall movement and prolonged uninterrupted sitting do not necessarily occur together. Although both groups accumulated relatively low levels of physical activity, the prolonged sedentary phenotype was characterised by substantially greater accumulation of sitting in extended bouts and fewer interruptions to sedentary time. This finding suggests that sedentary accumulation and physical inactivity represent related but separate dimensions of movement behaviour.

These phenotypes likely reflect underlying differences in daily routines and environmental contexts. For example, occupation is a key determinant of movement behaviour, with individuals in predominantly sitting occupations accumulating more sedentary time and fewer breaks than those in more active roles (Clemes et al., 2014). Such patterns may extend beyond working hours, suggesting that movement behaviours may be influenced by broader behavioural routines rather than specific domains alone. This provides a plausible explanation for the prolonged sedentary phenotype observed in the present study, characterised by extended sitting and reduced movement fragmentation. These findings are consistent with emerging work demonstrating that individuals differ not only in overall activity levels but also in the way movement behaviours are accumulated throughout the day.

The predominance of sedentary behaviour and low levels of MVPA observed in this study are consistent with behavioural patterns associated with increased risk of cancer, cardiometabolic disease, and premature mortality. Higher levels of physical activity have been robustly associated with reduced risk across multiple cancer sites and major chronic diseases, with dose–response relationships observed in large pooled analyses (Matthews et al., 2020; Moore et al., 2016). Conversely, higher levels of sedentary behaviour have been linked with a range of adverse health outcomes, including colorectal, endometrial and ovarian cancer, cardiovascular disease, type 2 diabetes, and all-cause mortality (Swain et al., 2023; Ekelund et al., 2019; Saint-Maurice et al., 2020). The higher sedentary time and lower activity levels observed among individuals with higher BMI are also consistent with established evidence linking adiposity, physical inactivity, and increased risk of both cancer and cardiometabolic disease (Lauby-Secretan et al., 2016). The behavioural profile observed in this cohort, characterised by high sedentary time, limited MVPA, and substantial time accumulated in prolonged sedentary bouts, may therefore represent a movement profile associated with elevated chronic disease risk. Although differences across demographic subgroups were modest, even small shifts in activity distribution may have meaningful implications at the population level.

### Strengths and limitations

This study has several important strengths. The relatively large sample size and good wear-time compliance support the reliability of the findings, and the use of device-based measures reduced biases associated with self-reported data. A key strength was the use of complementary posture- and intensity-based accelerometers, providing a more complete characterisation of movement behaviours through assessment of postural allocation, sedentary accumulation, movement transitions, stepping behaviour, and physical activity intensity. In addition, the study examined multiple aspects of movement behaviour, including overall activity levels, intensity, and accumulation patterns. The identification of behavioural phenotypes is a further strength, highlighting meaningful differences in how individuals accumulate movement behaviours that are not captured by traditional subgroup analyses.

Several limitations should be considered. Although a 7-day monitoring period is commonly used, it may not fully reflect longer-term habitual behaviour, nor does it capture changes over time that may be important predictors of health outcomes. In addition, participants were included if they provided at least one valid wear day from both devices. While this criterion was selected to maximise sample retention and preserve variability for this descriptive analysis, a single day of monitoring may not fully capture usual movement behaviours at the individual level. As with many cohort studies, participants may not be fully representative of the wider population. While device-based measurement improves accuracy, some activities such as water-based activities or cycling, may not be well captured, and short periods of non-wear may lead to small underestimation of activity. Differences between devices may also affect estimates, particularly for sedentary time. In addition, accelerometers do not provide information on the context in which behaviours occur (e.g. work, transport, or leisure), which may be important for understanding behavioural patterns.

## Conclusions

In this large cohort study of middle-aged and older Australian adults, device-measured movement behaviours were characterised by a predominance of sedentary and light-intensity activity, low levels of moderate-to-vigorous physical activity, and substantial variability between individuals. Differences across demographic groups were modest relative to this heterogeneity. The identification of distinct behavioural phenotypes, particularly the separation of low-activity and prolonged sedentary profiles, suggests that movement behaviours may be better understood by considering multiple behavioural characteristics rather than activity volume alone. Together, these findings emphasise the importance of considering both the volume and pattern of movement behaviours when characterising population health and designing interventions targeting movement patterns relevant to chronic disease prevention.

## Data Availability

The data underlying this article cannot be shared publicly due to compliance with participant consent and existing ethics approvals. They can be made available on reasonable request, with appropriate ethics approval, to.

## Acknowledgments and Funding

The Australian Breakthrough Cancer (ABC) Study cohort recruitment was funded by Cancer Council Victoria and a generous gift from the Geary Estate. Ongoing data and sample collection and participant engagement is supported by Council Cancer Victoria. Gandel Philanthropy provided support for the collection of accelerometer data. The collection of accelerometer data was also supported by the Thompson Estate. Dr Verswijveren was supported by a Deakin University Postdoctoral Research Fellow.

The ABC Study was conceptualised and established under the leadership of Professor Graham G. Giles, AM. We thank the ABC Study team and the tens of thousands of ABC Study participants from across Australia who generously contribute their time, data and samples to make the study possible.

## Author Contribution

Jay Keatley: Conceptualization, data curation, formal analysis, investigation, visualization, methodology, writing–original draft, writing–review and editing.

Nga Nguyen: Data curation

Paddy C. Dempsey: Methodology, writing–review and editing.

Simone J.J.M. Verswijveren: Methodology, writing–review and editing.

Julie K. Bassett: writing–review and editing.

Roger L. Milne: Resources, funding acquisition, writing–review and editing.

Brigid M. Lynch: Conceptualization, supervision, resources, funding acquisition, investigation, methodology, project administration, writing–review and editing.

## Ethics Approval

The ABC Study has been approved by the Human Research Ethics Committee of Cancer Council Victoria. This study has been carried out according to the National Statement on Ethical Conduct in Human Research (2007).

