## Supplementary Material for "Distribution and Patterns of Device-Measured Movement Behaviours in middle-aged to older Australian adults: the ABC Accelerometer Sub-Study"

### Supplementary Methods

Device-specific quality-control procedures were applied independently to activPAL and ActiGraph datasets prior to analysis. Daily activPAL records were excluded based on device error flags and insufficient monitored time (<480 min/day). ActiGraph records were excluded based on insufficient wear time (<480 min/day), implausible activity allocation relative to wear time, and excessive vector magnitude unit (VMU) values.

Behaviour classifications were validated by comparing the sum of behaviour-specific durations with total monitored time. Potentially implausible values were identified using prespecified biologically plausible thresholds, including daily step counts <100 or >40,000 steps/day, sitting time outside 300–900 min/day, stepping time outside 30–180 min/day, and sit-to-stand transitions outside 20–120/day. These checks were used to assess data quality and inform sensitivity analyses. Days flagged for excessive VMU counts were excluded from the ActiGraph dataset.

Data integrity procedures included verification of participant identifiers, assessment of missing data, removal of duplicate records, and harmonisation of variables across datasets. Sensitivity analyses examined alternative inclusion criteria (≥3 valid days), exclusion of extreme values, and device-specific samples

### Supplementary Tables

**Supplementary Table 1a**: Daily movement behaviours measured by activPAL and actiGraph; overall and by BMI group

**Supplementary Table 1a: Daily movement behaviours measured by activPAL and ActiGraph, overall and by BMI group**

| Characteristic | Overall N = 3,352[1] | Normal (18.5–24.9) N = 1,161[1] | Obese (≥30) N = 809[1] | Overweight (25–29.9) N = 1,348[1] | Underweight (<18.5) N = 34[1] |
|---|---|---|---|---|---|
| activPAL sitting time, min/day | 378.8 (97.7) | 375.9 (94.6) | 388.2 (106.3) | 375.0 (94.6) | 400.9 (96.0) |
| activPAL standing time, min/day | 161.0 (65.4) | 169.1 (63.2) | 151.1 (67.1) | 160.3 (65.6) | 152.0 (55.1) |
| activPAL stepping time, min/day | 64.5 (26.8) | 70.5 (27.5) | 55.4 (24.1) | 64.9 (26.3) | 62.6 (28.4) |
| activPAL lying time, min/day | 282.0 (82.6) | 280.9 (77.9) | 284.9 (86.6) | 281.5 (84.5) | 263.6 (66.7) |
| activPAL steps/day | 5,192 (2,361) | 5,787 (2,515) | 4,346 (2,044) | 5,192 (2,245) | 4,941 (2,382) |
| ActiGraph sedentary time, min/day | 583.8 (128.2) | 576.8 (125.4) | 594.1 (136.1) | 583.3 (124.5) | 594.6 (159.4) |
| ActiGraph light activity, min/day | 290.9 (74.4) | 298.4 (71.6) | 273.1 (78.3) | 295.1 (72.7) | 295.8 (68.0) |
| ActiGraph moderate activity, min/day | 31.7 (23.4) | 36.8 (24.5) | 23.4 (19.4) | 32.4 (23.4) | 26.5 (17.1) |
| ActiGraph vigorous activity, min/day | 1.5 (4.6) | 2.5 (5.9) | 0.6 (3.0) | 1.2 (3.9) | 0.4 (1.1) |
| ActiGraph MVPA, min/day | 33.2 (25.0) | 39.3 (26.6) | 24.0 (20.3) | 33.6 (24.5) | 26.9 (17.4) |
| ActiGraph steps/day | 7,159 (5,303, 9,503) | 8,202 (6,213, 10,661) | 5,833 (4,142, 7,827) | 7,179 (5,425, 9,445) | 7,176 (5,225, 9,722) |

[1] Mean (SD); Median (Q1, Q3)

Values are presented as mean (SD), except steps/day which are presented as median (IQR). MVPA = moderate-to-vigorous physical activity.

**Supplementary 1b:** Daily movement behaviours measured by activPAL and actiGraph, overall, and by Age group

**Supplementary Table 1b: Daily movement behaviours measured by activPAL and ActiGraph, overall and by Age group**

| Characteristic | Overall N = 3,425[1] | 30-39 N = 4[1] | 40-49 N = 398[1] | 50-59 N = 1,104[1] | 60-69 N = 1,435[1] | 70-80 N = 484[1] |
|---|---|---|---|---|---|---|
| activPAL sedentary time, min/day | 379.3 (98.6) | 423.5 (103.9) | 370.1 (100.2) | 374.8 (101.1) | 380.5 (94.8) | 393.6 (100.9) |
| activPAL standing time, min/day | 160.8 (65.4) | 213.0 (39.7) | 159.8 (62.0) | 161.0 (65.4) | 160.2 (65.8) | 162.4 (67.2) |
| activPAL stepping time, min/day | 64.5 (26.8) | 77.6 (26.9) | 65.2 (25.8) | 65.1 (26.6) | 65.2 (27.4) | 60.1 (26.2) |
| activPAL lying time, min/day | 282.2 (83.0) | 360.2 (109.7) | 289.0 (86.0) | 279.6 (82.9) | 280.8 (82.9) | 286.2 (80.5) |
| activPAL steps/day | 5,184 (2,359) | 6,037 (2,040) | 5,222 (2,169) | 5,291 (2,360) | 5,234 (2,424) | 4,758 (2,275) |
| ActiGraph sedentary time, min/day | 584.0 (127.9) | 535.8 (112.6) | 586.0 (123.9) | 587.0 (130.7) | 577.1 (125.3) | 596.1 (131.2) |
| ActiGraph light activity, min/day | 290.4 (74.3) | 288.6 (118.7) | 293.5 (70.1) | 294.3 (76.1) | 291.7 (74.3) | 275.6 (72.0) |
| ActiGraph moderate activity, min/day | 31.6 (23.4) | 29.0 (21.6) | 32.8 (19.5) | 34.4 (22.9) | 31.3 (23.8) | 25.2 (25.1) |
| ActiGraph vigorous activity, min/day | 1.5 (4.5) | 1.0 (1.4) | 2.6 (5.8) | 2.0 (5.1) | 1.2 (4.1) | 0.4 (2.2) |
| ActiGraph MVPA, min/day | 33.1 (24.9) | 30.0 (22.8) | 35.3 (22.0) | 36.4 (24.6) | 32.4 (25.0) | 25.6 (26.0) |
| ActiGraph steps/day | 7,140 (5,291, 9,478) | 7,497 (4,917, 9,983) | 7,402 (5,720, 9,600) | 7,553 (5,605, 9,890) | 7,175 (5,207, 9,457) | 5,992 (4,629, 8,341) |

[1] Mean (SD); Median (Q1, Q3)

Values are presented as mean (SD), except steps/day which are presented as median (IQR). MVPA = moderate-to-vigorous physical activity.

**Supplementary 1c:** Daily movement behaviours measured by activPAL and actiGraph, overall, and by Education

**Supplementary Table 1c: Daily movement behaviours measured by activPAL and ActiGraph, overall and by Education**

| Characteristic | Overall<br>N = 3,406† | Certificate/diploma<br>N = 976† | Junior Secondary<br>N = 410† | Primary Education or less<br>N = 20† | Senior Secondary Education<br>N = 346† | University degree or higher<br>N = 1,654† |
|---|---|---|---|---|---|---|
| **activPAL sedentary time, min/day** | 379.3 (98.7) | 376.3 (99.6) | 374.0 (103.6) | 388.8 (92.7) | 383.7 (98.4) | 381.2 (97.0) |
| **activPAL standing time, min/day** | 160.7 (65.3) | 163.8 (64.9) | 166.7 (74.5) | 159.6 (82.2) | 155.9 (61.0) | 158.5 (63.7) |
| **activPAL stepping time, min/day** | 64.5 (26.8) | 63.4 (26.5) | 62.8 (27.8) | 61.0 (24.9) | 63.5 (26.4) | 65.7 (26.8) |
| **activPAL lying time, min/day** | 282.2 (82.9) | 281.0 (81.3) | 284.3 (89.2) | 325.7 (131.0) | 281.4 (81.3) | 281.9 (81.8) |
| **activPAL steps/day** | 5,187 (2,360) | 5,011 (2,259) | 4,908 (2,391) | 4,729 (2,177) | 5,137 (2,432) | 5,377 (2,383) |
| **ActiGraph sedentary time, min/day** | 583.8 (127.7) | 584.4 (134.1) | 580.2 (135.6) | 570.4 (97.5) | 583.8 (129.7) | 584.5 (121.7) |
| **ActiGraph light activity, min/day** | 290.4 (74.3) | 296.9 (75.9) | 298.2 (80.9) | 268.4 (62.2) | 288.6 (75.2) | 285.4 (71.1) |
| **ActiGraph moderate activity, min/day** | 31.6 (23.4) | 29.8 (22.4) | 25.7 (23.4) | 20.6 (21.8) | 31.3 (23.8) | 34.4 (23.6) |
| **ActiGraph vigorous activity, min/day** | 1.5 (4.5) | 1.3 (4.2) | 0.5 (1.9) | 0.1 (0.4) | 1.4 (4.7) | 1.9 (5.1) |
| **ActiGraph MVPA, min/day** | 33.1 (25.0) | 31.1 (24.2) | 26.2 (23.7) | 20.7 (21.9) | 32.7 (25.1) | 36.3 (25.2) |
| **ActiGraph steps/day** | 7,141 (5,302, 9,494) | 7,025 (5,153, 9,401) | 6,511 (4,592, 9,012) | 6,087 (3,640, 8,325) | 7,314 (5,324, 9,441) | 7,435 (5,521, 9,678) |

† Mean (SD); Median (Q1, Q3)

Values are presented as mean (SD), except steps/day which are presented as median (IQR). MVPA = moderate-to-vigorous physical activity.

**Supplementary Table 2a** Sedentary accumulation patterns measured by activPAL, overall, and by BMI group

**Supplementary Table 2a: Sedentary accumulation patterns measured by activPAL, overall and by BMI group**

| Characteristic | Overall<br>N = 3,352† | Normal (18.5–24.9)<br>N = 1,161† | Obese (≥30)<br>N = 809† | Overweight (25–29.9)<br>N = 1,348† | Underweight (<18.5)<br>N = 34† |
|---|---|---|---|---|---|
| **Total sitting time, min/day** | 378.8 (97.7) | 375.9 (94.6) | 388.2 (106.3) | 375.0 (94.6) | 400.9 (96.0) |
| **Sit-to-stand transitions, n/day** | 27.2 (9.3) | 28.5 (9.7) | 25.1 (8.8) | 27.1 (9.1) | 29.9 (10.2) |
| **Stand-to-sit transitions, n/day** | 27.1 (9.3) | 28.5 (9.7) | 25.1 (8.8) | 27.1 (9.1) | 29.9 (10.2) |
| **Sedentary bouts ≥30 min, n/day** | 2.9 (1.0) | 2.7 (1.0) | 3.1 (1.1) | 2.9 (1.0) | 2.8 (0.8) |
| **Sedentary bouts ≥60 min, n/day** | 1.1 (0.5) | 1.0 (0.5) | 1.2 (0.6) | 1.0 (0.5) | 1.0 (0.5) |
| **Time in sedentary bouts ≥30 min, min/day** | 254.7 (92.7) | 250.0 (90.3) | 270.1 (98.9) | 249.3 (89.7) | 267.2 (97.9) |
| **Time in sedentary bouts ≥60 min, min/day** | 179.9 (84.9) | 179.4 (83.8) | 190.5 (89.4) | 173.6 (82.3) | 193.8 (92.4) |

† Mean (SD)

**Supplementary Table 2b** Sedentary accumulation patterns measured by activPAL, overall and by Age group

**Supplementary Table 2b: Sedentary accumulation patterns measured by activPAL, overall and by Age Group**

| Characteristic | Overall N = 3,421[1] | 40-49 N = 398[1] | 50-59 N = 1,104[1] | 60-69 N = 1,435[1] | 70-80 N = 484[1] |
|---|---|---|---|---|---|
| **Total sedentary time, min/day** | 379.3 (98.6) | 370.1 (100.2) | 374.8 (101.1) | 380.5 (94.8) | 393.6 (100.9) |
| **Sit-to-stand transitions, n/day** | 27.1 (9.4) | 28.8 (9.2) | 27.3 (9.4) | 27.0 (9.6) | 25.7 (8.2) |
| **Stand-to-sit transitions, n/day** | 27.1 (9.4) | 28.8 (9.2) | 27.3 (9.4) | 27.0 (9.6) | 25.7 (8.2) |
| **Sedentary bouts ≥30 min, n/day** | 2.9 (1.0) | 2.7 (1.0) | 2.8 (1.0) | 2.9 (1.0) | 3.1 (1.1) |
| **Sedentary bouts ≥60 min, n/day** | 1.1 (0.5) | 1.0 (0.5) | 1.0 (0.5) | 1.1 (0.5) | 1.2 (0.6) |
| **Time in sedentary bouts ≥30 min, min/day** | 255.2 (93.7) | 243.7 (95.3) | 251.2 (95.7) | 256.4 (91.2) | 270.2 (93.1) |
| **Time in sedentary bouts ≥60 min, min/day** | 180.2 (86.0) | 169.9 (85.5) | 179.2 (88.9) | 180.4 (84.8) | 190.2 (82.5) |

[1] Mean (SD)

**Supplementary Table 2c** Sedentary accumulation patterns measured by activPAL, overall, and by Education

**Supplementary Table 2c: Sedentary accumulation patterns measured by activPAL, overall and by Education**

| Characteristic | Overall N = 3,405[1] | Certificate/diploma N = 976[1] | Junior Secondary N = 410[1] | Primary Education or less N = 20[1] | Senior Secondary Education N = 346[1] | University degree or higher N = 1,653[1] |
|---|---|---|---|---|---|---|
| **Total sedentary time, min/day** | 379.3 (98.7) | 376.3 (99.6) | 374.0 (103.6) | 388.8 (92.7) | 383.7 (98.4) | 381.3 (97.0) |
| **Sit-to-stand transitions, n/day** | 27.1 (9.3) | 27.0 (9.4) | 27.0 (10.2) | 26.2 (9.5) | 26.8 (9.0) | 27.3 (9.2) |
| **Stand-to-sit transitions, n/day** | 27.1 (9.3) | 27.0 (9.3) | 27.0 (10.2) | 26.1 (9.4) | 26.8 (9.0) | 27.3 (9.2) |
| **Sedentary bouts ≥30 min, n/day** | 2.9 (1.0) | 2.8 (1.1) | 2.8 (1.1) | 3.2 (1.1) | 2.9 (1.1) | 2.9 (1.0) |
| **Sedentary bouts ≥60 min, n/day** | 1.1 (0.5) | 1.0 (0.5) | 1.1 (0.6) | 1.3 (0.6) | 1.1 (0.5) | 1.1 (0.5) |
| **Time in sedentary bouts ≥30 min, min/day** | 255.2 (93.8) | 252.8 (93.5) | 250.2 (97.1) | 269.1 (94.0) | 261.2 (96.9) | 256.4 (92.5) |
| **Time in sedentary bouts ≥60 min, min/day** | 180.1 (86.1) | 178.0 (85.8) | 178.0 (86.4) | 189.0 (91.7) | 186.0 (89.5) | 180.6 (85.5) |

[1] Mean (SD)

**Supplementary Table 4:** Behavioural phenotype clusters and device-measured movement characteristics within each cluster

**Supplementary Table 4. Behavioural phenotype clusters and device-measured movement characteristics within each cluster**

| Movement characteristic | Active/Fragmented N = 803[†] | Low Activity N = 1,406[†] | Prolonged Sedentary N = 1,216[†] |
|---|---|---|---|
| **Steps/day** | 8,020.3 (2,397.3) | 4,505.8 (1,471.1) | 4,096.2 (1,535.6) |
| **Light activity, min/day** | 331.4 (72.3) | 303.6 (66.7) | 248.2 (62.3) |
| **Moderate activity, min/day** | 59.0 (25.0) | 22.9 (13.4) | 23.6 (16.5) |
| **Vigorous activity, min/day** | 4.2 (7.9) | 0.6 (1.9) | 0.7 (2.3) |
| **MVPA, min/day** | 63.3 (26.1) | 23.4 (13.8) | 24.4 (17.2) |
| **Sedentary time, min/day** | 363.6 (80.9) | 314.6 (68.0) | 464.6 (73.7) |
| **Standing time, min/day** | 186.5 (69.8) | 168.0 (68.2) | 135.5 (48.3) |
| **Stepping time, min/day** | 95.7 (27.1) | 57.9 (18.1) | 51.3 (17.3) |
| **Lying time, min/day** | 299.3 (92.6) | 279.3 (85.0) | 274.4 (71.7) |
| **Sit-to-stand transitions, n/day** | 33.4 (11.6) | 25.6 (7.9) | 24.7 (7.2) |
| **Sedentary time ≥30 min bouts, min/day** | 219.0 (74.1) | 196.5 (53.4) | 347.0 (67.2) |
| **Sedentary time ≥60 min bouts, min/day** | 147.5 (71.3) | 129.5 (49.0) | 260.4 (67.9) |

[†] Mean (SD)

**Supplementary Table 5:** Sensitivity analysis summary across analytic samples

**Supplementary Table 5. Sensitivity analysis summary across analytic samples**

Descriptive summaries for primary, stricter wear-time, outlier-excluded, and device-specific samples

| Sensitivity dataset | N participants | N records | Sedentary, mean (SD) | Standing/Light, mean (SD) | Stepping/MVPA, mean (SD) | Steps, mean (SD) |
|---|---|---|---|---|---|---|
| activPAL | | | | | | |
| Primary activPAL | 3428 | 56596 | 377.4 (272.3) | 155.8 (159.2) | 62.0 (68.1) | 4977.1 (5731.4) |
| activPAL >=3 valid days | 3281 | 55351 | 380.0 (271.9) | 156.5 (159.3) | 62.2 (68.1) | 4995.5 (5734.3) |
| activPAL outliers removed | 3428 | 55836 | 377.9 (273.7) | 154.0 (159.1) | 59.2 (64.1) | 4715.7 (5282.2) |
| All activPAL participants | 3450 | 56962 | 377.0 (272.2) | 155.7 (159.2) | 62.0 (68.1) | 4975.7 (5731.9) |
| ActiGraph | | | | | | |
| Primary ActiGraph | 3618 | 24814 | 584.5 (164.7) | 291.4 (101.3) | 33.2 (33.9) | 19646.9 (88662.5) |
| ActiGraph >=3 valid days | 3589 | 24756 | 584.7 (164.7) | 291.6 (101.2) | 33.2 (33.9) | 19682.9 (88763.1) |
| ActiGraph outliers removed | 3613 | 24325 | 588.4 (161.8) | 289.5 (100.2) | 32.6 (33.3) | 12024.6 (42658.9) |
| All ActiGraph participants | 3859 | 26409 | 583.9 (164.3) | 291.3 (101.6) | 33.2 (34.1) | 19079.5 (86565.0) |

Values are presented as mean (SD). For activPAL, the second and third behaviour columns correspond to standing and stepping, respectively. For ActiGraph, they correspond to light-intensity activity and MVPA.

### Supplementary Figures

**Supplementary Figure 1a:** Distributions of sedentary time and activity intensities, by BMI group

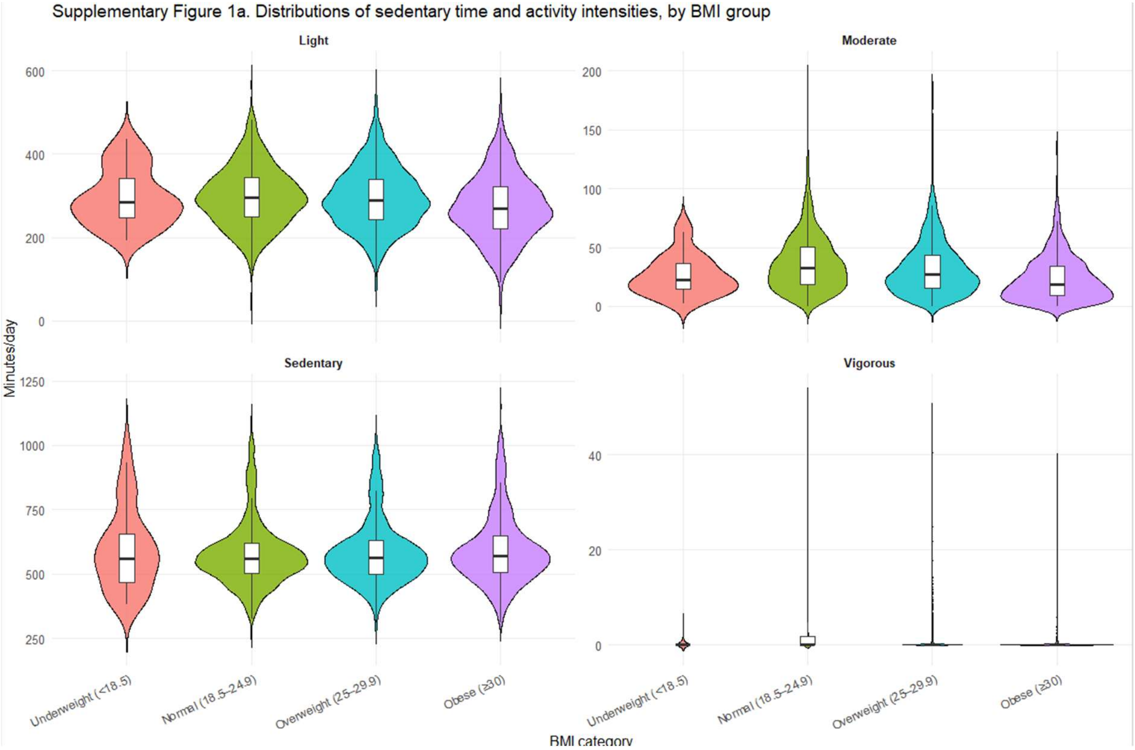

**Supplementary Figure 1b:** Distributions of sedentary time and activity intensities, by Age group

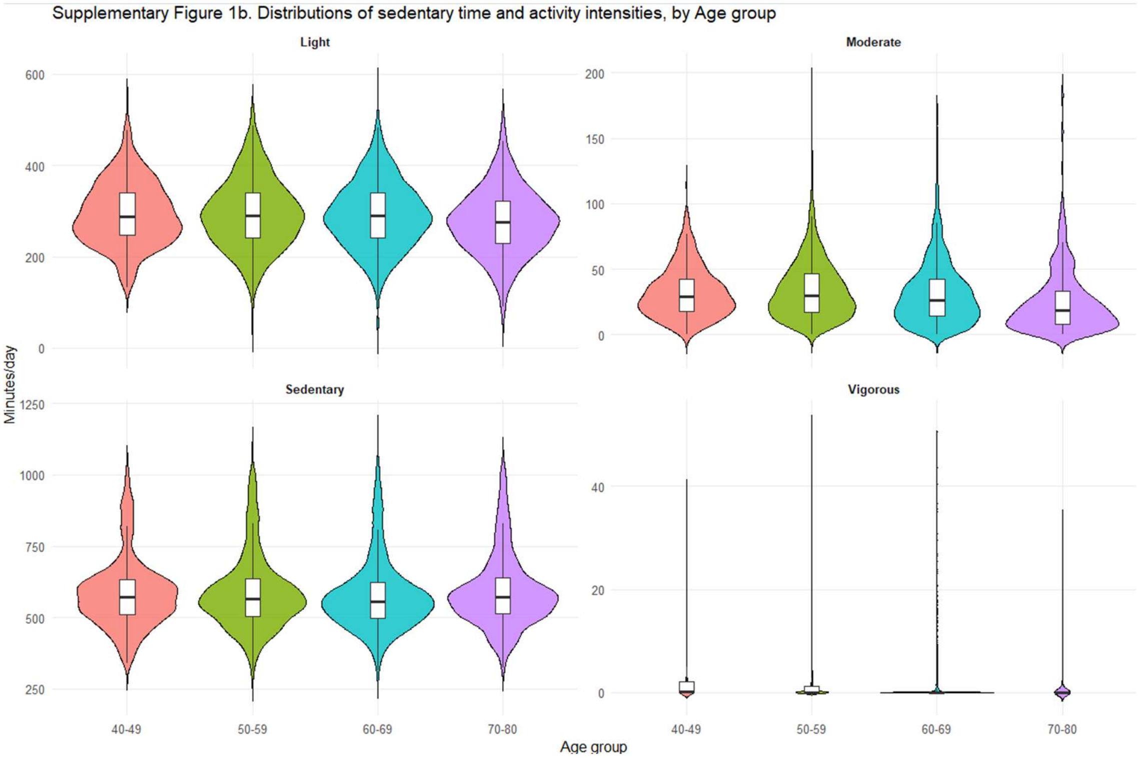


**Supplementary Figure 1c.** Distributions of sedentary time and activity intensities, by Education

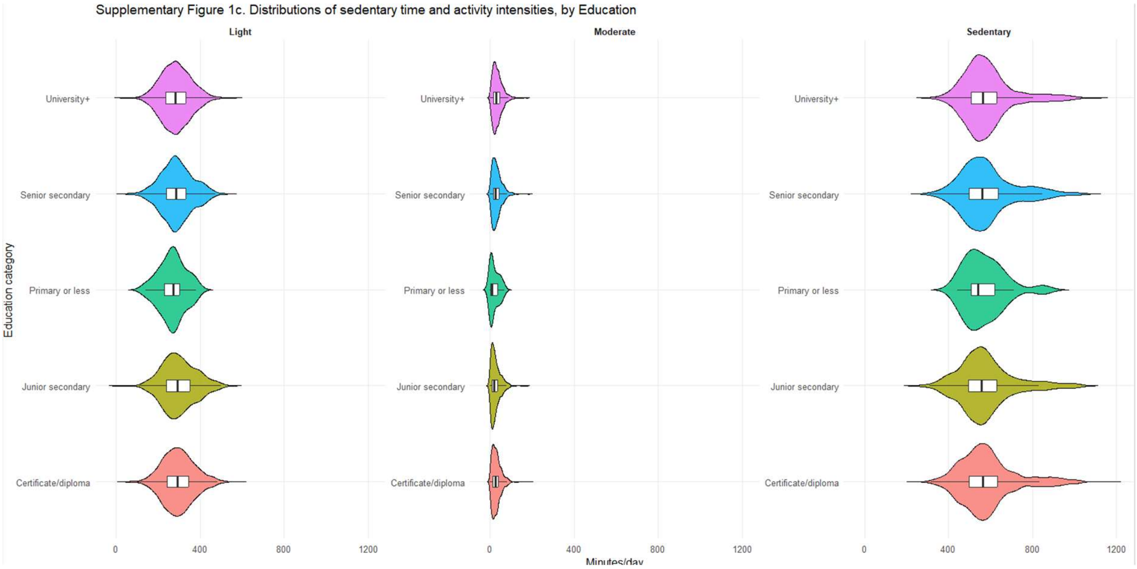

**Supplementary Figure 2a.** Distribution of step counts, cadence, by BMI group

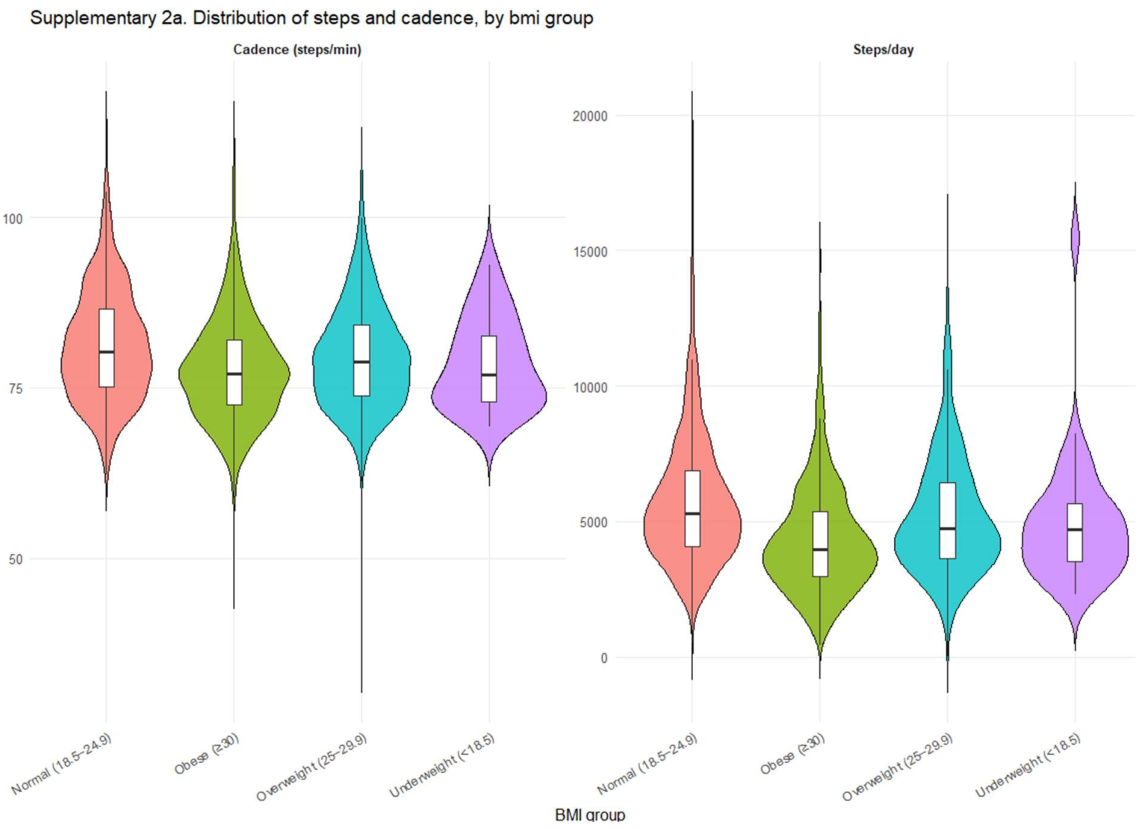

**Supplementary Figure 2b.** Distribution of step counts, cadence, by Age group

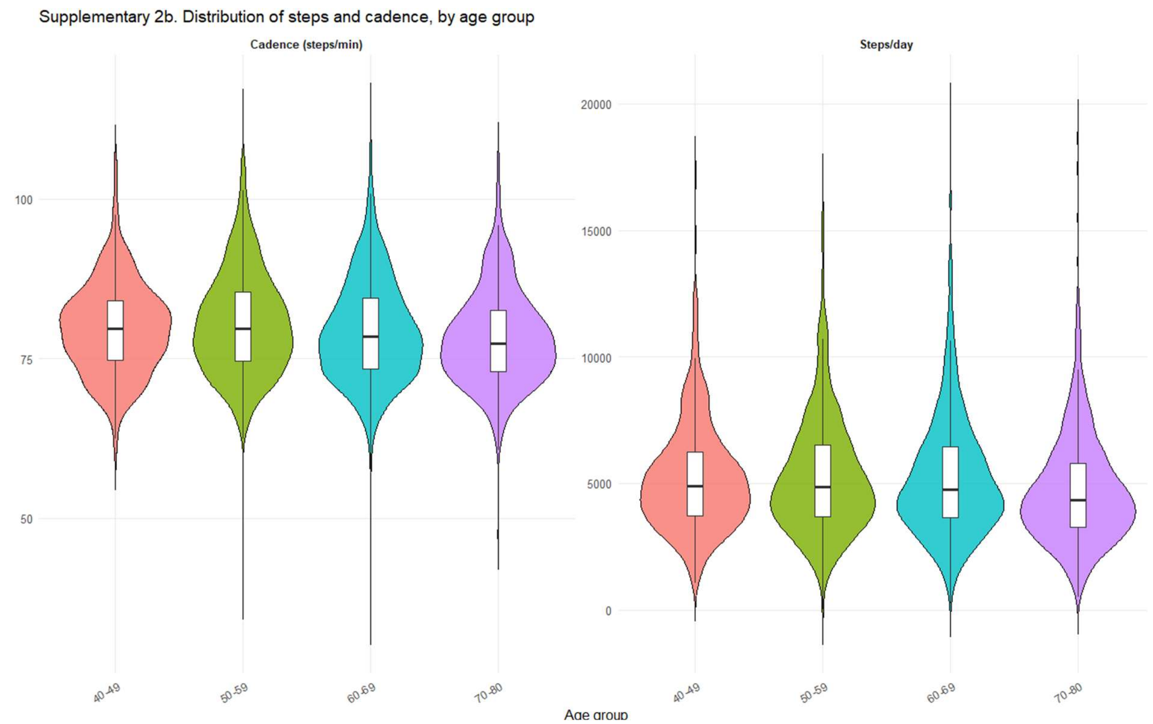


**Supplementary Figure 2c.** Distribution of step counts, cadence, by Education

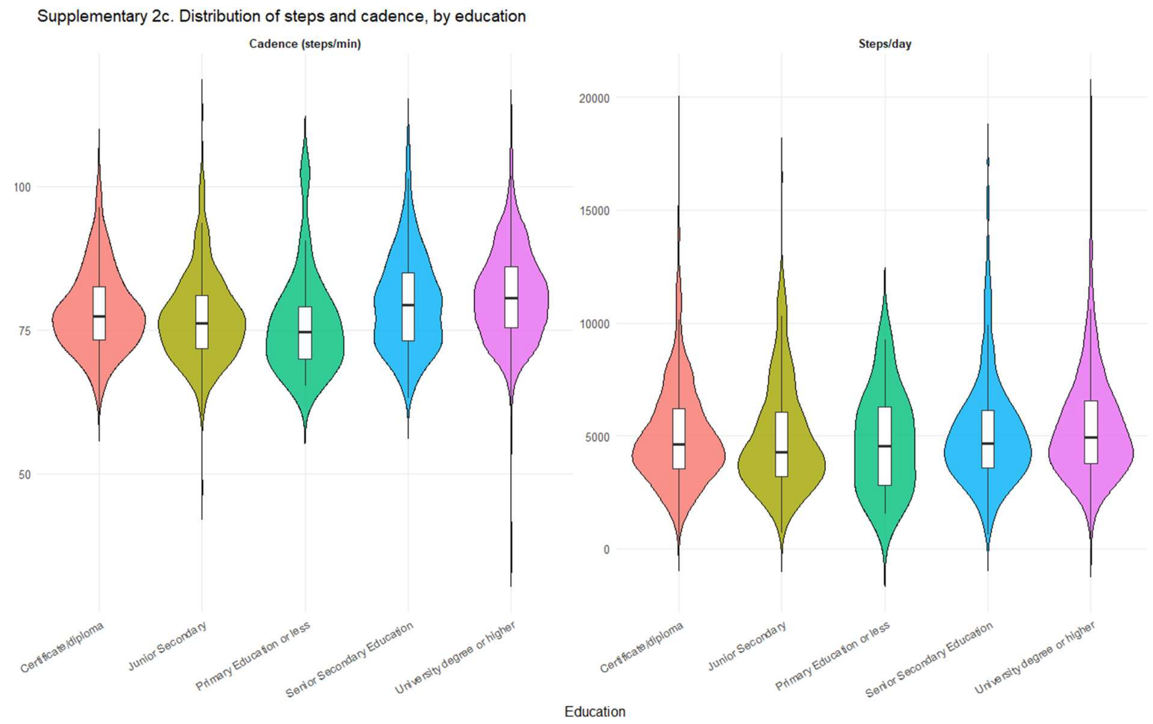

**Supplementary Figure 3:** Cluster heatmap of behavioural phenotypes

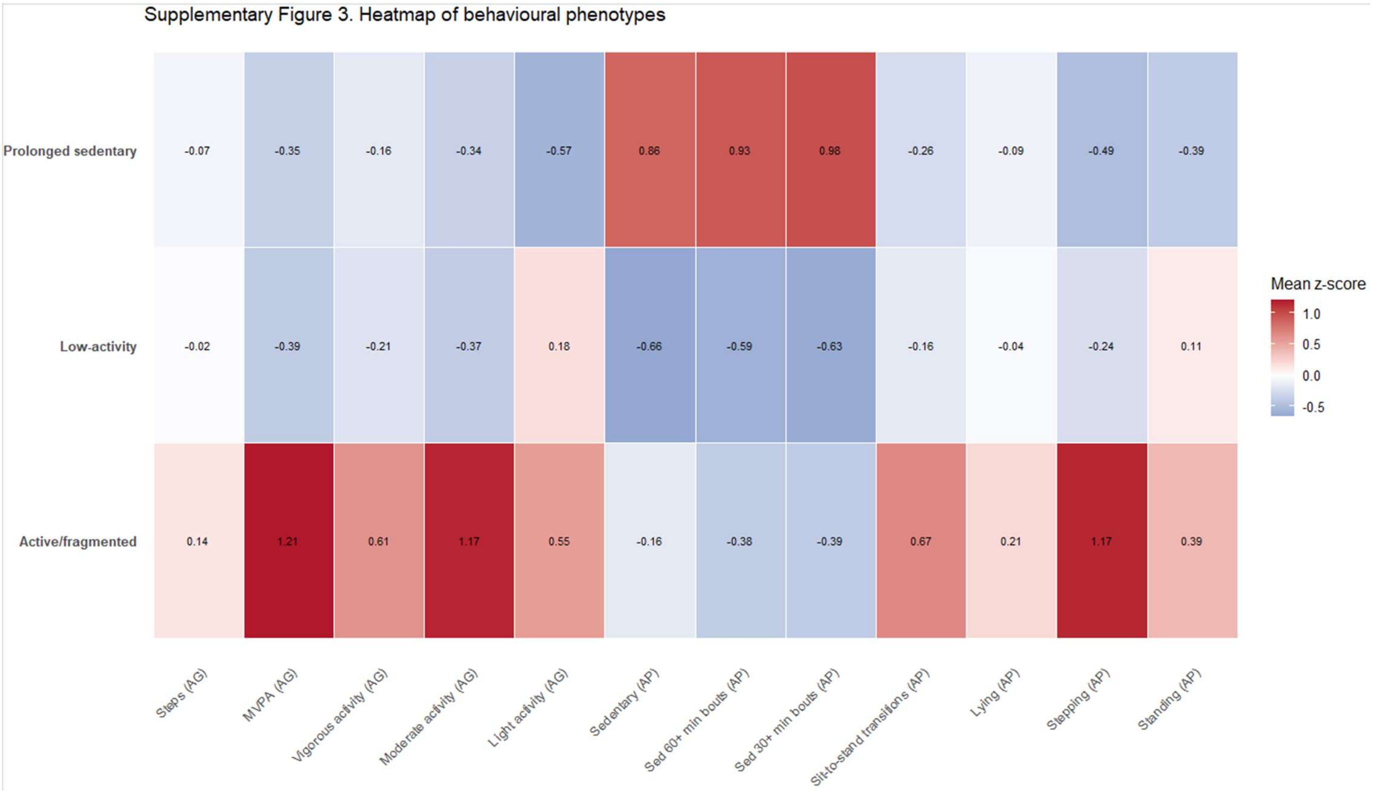


*Rows represent behavioural phenotypes identified using k-means clustering and columns represent activPAL-derived posture measures and ActiGraph-derived intensity measures. Values are mean z-scores relative to the cohort mean (0 = average), with red indicating higher-than-average values and blue indicating lower-than-average values. Active/fragmented phenotype (Cluster 1) showed higher activity levels and movement fragmentation), Low activity (Cluster 2) showed lower overall activity), and Prolonged Sedentary (Cluster 3) showed greater sedentary accumulation.*
